# High risk of infection caused Posttraumatic Stress symptoms in individuals with poor sleep quality: A study on influence of Coronavirus disease (COVID-19) in China

**DOI:** 10.1101/2020.03.22.20034504

**Authors:** Fan Zhang, Zhilei Shang, Haiying Ma, Yanpu Jia, Luna Sun, Xin Guo, Lili Wu, Zhuoer Sun, Yaoguang Zhou, Yan Wang, Nianqi Liu, Weizhi Liu

**Author notes:** these authors contributed equally to this work (Fan Zhang, Zhilei Shang, Haiying Ma, Yanpu Jia).

## Abstract

The influence of the outbreak of coronavirus disease (COVID-19) on mental health was poorly understood. The present study aimed to exam sleep problems and posttraumatic stress symptoms (PTSS) in Chinese immediately after the massive outbreak of COVID-19. A total of 2027 Chinese participated in the present study. Wuhan-expose history, sleep quality and PTSS were measured with self-rating scales. Results showed that **t**here were significant differences of PCL-5 and of sleep quality scores in different data-collection dates (*p*s<0.05). There were significant differences of PCL-5 scores (*t*=-2.93, *p*<0.05) and latency onset of sleep (χ2=9.77, *p*<0.05) between participants with and without Wuhan-expose history. The interaction effect of Wuhan exposure history× sleep quality significantly influenced PCL-5 (*p*s<0.05). These results indicate that keeping good sleep quality in individuals with high infectious risk is a way to prevent PTSS.

## Introduction

On Dec 31, 2019, the World Health Organization (WHO) China Office was informed of cases of an unknown etiology detected in Wuhan City, Hubei province of China (WHO, 2020a). On Jan 7, 2020, a novel coronavirus, which had not been documented before was identified by the Chinese Center for Disease Control and Prevention (CDC) from the throat swab sample of a patient, and was confirmed as the cause of the Coronavirus disease (COVID-19) by WHO (2020a). On Jan 30, 2020 the outbreak of COVID-19 has been determined by the WHO as a Public Health Emergency of International Concern (PHEIC) (WHO, 2020b). Thus far (up to Feb 4, 2020), more than 20,000 firmed cases have been confirmed in China, and more than 20 countries reported confirmed cases (WHO, 2020c).

According to the Diagnostic and Statistical Manual of Mental Disorders-5th edition (DSM-5) (APA, 2013), Posttraumatic Stress Disorder (PTSD) is characterized by four symptom groups: (a) involuntary memories of the trauma such as intrusions or nightmares; (b) persistent avoidance of stimuli associated with the traumatic event; (c) negative alterations in cognitions and mood that are associated with the trauma; and (d) alterations in arousal and reactivity that are associated with the trauma. Also in DSM-5, exposure to traumatic events is the immediate causes of PTSD and an essential condition of diagnosis. Being infected with a life-threatening physical illness is a traumatic event and can lead to posttraumatic stress symptoms. For example, 12%-15% survivors of SARS showed one type of PTSD symptom (PTSS) at one month after discharge (Wu et al, 2005); the morbidity of PTSD in patients with traumatic injury reached 9.6% at months (Bryant et al., 2017). In addition, not only being attacked by traumatic illness could cause PTSD symptoms, being contacts and carers of a deadly virus could also lead to symptoms like alternations in mood and arousal (Tine et al., 2016). Living in the endemic area, every Chinese is a carer and a (possible) contact of COVID-19. Although having experienced the outbreak of Severe Acute Respiratory Syndrome (SARS), a severe infectious disease which had caused hundreds of death in China in 2003 (WHO, 2003), the traumatic course of the infection and the characteristic of human transmission of novel coronavirus (Li et al., 2020) were beyond expectation, causing fear, helplessness and PTSS among Chinese people. Hence in the present study, we focused on the acute PTSS in Chinese under the background of massive COVID-19 outbreak.

Previous studies have proved that massive outbreaks of epidemics like SARS and Ebola virus disease (EVD) could cause sleep problems in relevant individuals. For example, Yu et al. (2005) reported that midlife women felt “sleep was restless” during SARS and this problem was closely associated with their overall negative emotions. Another study showed that chronic post-SARS symptoms were characterized by sleep-related problems (Moldofsky and Patcai, 2011). Survivors of EVD also reported various sleep quality problems (Keita et al., 2017). More importantly, Sleep and PTSS are closely related, and sleep disturbances are core features of PTSD. According to the model raised by Richard et al. (2020), sleep disturbances (short sleep duration, frequent arousals, and et al.) could lead to maladaptive sleep-related compensatory behaviors and cause hyperarousal and anxiety-related disorders like PTSD. Sleep disruption also promoted anxious emotions by disrupt emotion regulation and adaptive memory functions. Thus, it was reasonable to hypothesize that wide-spread transmission of COVID-19 could causes sleep problems in Chinese and result in PTSS.

In early December 2019, the first pneumonia case was identified in Wuhan, the capital city of Hubei province (Huang et al., 2020). With the spreading of the epidemic, Wuhan became the worst influenced area, where the mortality and incidence rate of COVID-19 were above the national level according to the National Health Commission of China (2020). Moreover, not only Wuhan residents were more likely to be infected, 72.3% nonresidents of Wuhan had contact with individuals lived in Wuhan (Guan et al., 2020). Wuhan expose history had become an important diagnosis standard and a source of fear. As a result, we collected data of Wuhan exposure to measure the risk of infection. We hypothesized that Wuhan residents and individuals who had been to Wuhan or contacted with Wuhan residents, would show more sleep problems and PTSS for they were faced with high possibility of infection of COVID-19. We also explored the relationship among the infection risk, sleep and PTSS to identify the role of sleep problems in the development of PTSD. We hope the current study could reflect the influence of COVID-19 on psychological well-being in Chinses and shed light on post-epidemic psychological intervention.

## Methods

The methods and reporting procedures were in accordance with the STROBE (STrengthening the Reporting of OBservational studies in Epidemiology) checklist.

### Participants

A total of 2032 subjects form 31 provinces in the mainland of China, Hong Kong, Macao and Taiwan participated in the present study. Criteria for inclusion was that subjects should be Chinese race and age ≥ 18 years at the time of the COVID-19 infection. Table 1 demonstrated demographic statistics of subjects. The average age of all subjects was 35.47±11.32.

**Table 1.** Demographic Statistics,exposure history and sleep status of all subjects.

|  | N | % |
| --- | --- | --- |
| **Gender** |  |  |
| Male | 786 | 38.8 |
| Female | 1241 | 61.2 |
| **Education Level** |  |  |
| High school or below | 254 | 12.5 |
| University or college | 1310 | 64.6 |
| Postgraduate or above | 463 | 22.8 |
| **Exposure history of Wuhan** |  |  |
| No | 1797 | 88.7 |
| Yes | 230 | 11.3 |
| **Subjective sleep quality** |  |  |
| Very good | 816 | 40.3 |
| Good | 859 | 42.4 |
| Poor | 320 | 15.8 |
| Very poor | 32 | 1.6 |
| **Latency onset of sleep** |  |  |
| No | 1358 | 67.0 |
| <Once a week | 312 | 15.4 |
| once or twice a week | 251 | 12.4 |
| >=Three times a week | 106 | 5.2 |
| **Sleep fragment** |  |  |
| No | 1133 | 55.9 |
| <Once a week | 312 | 15.4 |
| once or twice a week | 368 | 18.2 |
| >=Three times a week | 214 | 10.6 |
| **Sleep duration** |  |  |
| >7h | 996 | 49.1 |
| 6-7h | 645 | 31.8 |
| 5-6h | 314 | 15.5 |
| <5h | 72 | 3.6 |
| **Total** | 2027 | 100.0 |

All the study participants were provided with written informed consent. This project was approved by the Ethics Committee of the Naval Military Medical University.

### Wuhan expose history

Subjects who were Wuhan residents, or had been to Wuhan, or had contacted with anyone from Wuhan during the outbreak of COVID-19 scored 1 point and those not scored 0. A total of 230 subjects showed a history of exposure to Wuhan.

### Sleep quality

According to previous findings, certain types of sleep quality disturbances were more common across anxiety-related disorders than others (Leskin et al., 2002; Neylan, 1998). As a result, we only selected the most related items from the Pittsburgh Sleep Quality Index (PSQI) (Buysse et al., 1989) to investigate sleep quality. 4 items form PSQI were used to measure subject’s sleep quality. The first was the subjective sleep quality measuring participants’ satisfaction with sleep. Subject scored 0 if they had very good subjective sleep quality and scored 3 for very bad subjective sleep quality. The second item collected data on latency onset of sleep. Scores varied from 0∼3 with 0 standing for having no problems falling asleep and 3 standing for hard to fall asleep for 3 times or more within a week. The third item was the sleep fragment. Subjects score 1 for no occurrence of the problem and scored 3 if they had the problem for 3 times or more within a week. The last item collected data on everyday sleep duration and the scores varied form 0 (more than 7 hours) to 3 (less than 5 hours). In summary, 0 and 1 point of the 1^st^ item indicated very good and good subjective sleep quality and 2 and 3 indicated sleep quality problems. For the remaining items, 0 point indicated good sleep quality and 1 to 3 points indicated increasingly sever sleep quality problems.

### PTSS

PTSD Checklist for DSM-5 (PCL-5) (Weathers et al., 2013) was used to measure PTSS. PCL-5 is a self-report 5-point Likert-Type scale (1=not at all, and 5=extremely) with 20 items measuring PTSS of instruction, avoidance, numbing and hyperarousal. The Chinese version of PCL-5 showed excellent consistency and convergent validity in previous studies (Li et al., 2018; Liu et al., 2014). Internal consistency (Cronbach’s α) of PCL-5 in the present study was 0.87.

### Data collection

The data was collected during Jan 30, 2020 to Feb 3, 2020 through an online survey platform https://www.wjx.cn/. To complete the survey, participants had to answer all the listed questions in a time limitation of 2 to 30 minutes. Answers with response time below 2 mins or above 30mins were invalid and would be deleted in the following analysis.

### Data analysis

5 participants were deleted because of illegal response time. Data from 2027 participants were analyzed. One-way variance analysis (ANOVA) and Least-Significant Difference (LSD) test were used to analyze the differences of PTSS during each day of survey. Chi-square test was applied to analyzed differences of sleep quality by time and Wuhan-exposed history. Scores of PTSS were compared between subjects with and without Wuhan exposure history using *t*-test. Interaction effects of sleep and Wuhan exposure history on PTSS was analyzed using two-way ANOVAs and simple effect analyses.

## Results

### General results of sleep quality and PTSS

Figure 1 showed trends of PCL-5 scores and sleep quality during the 5-day survey. Both PCL-5 scores and sleep quality scores showed raising tendency from Jan 30, 2020 to Feb 2, 2020 and decreased in Feb 3, 2020. One-way ANOVA showed that there was a significant difference of PCL-5 scores on different dates, *F*=4.38, *p*<0.05. LSD test showed significant increases was illustrated from Jan 31 to Feb 2 (*p*s<0.05); however, the PCL-5 score in Feb 3, 2020 were significantly lower than that in Feb 2, 2020 (*p*<0.05).

**Figure 1.**
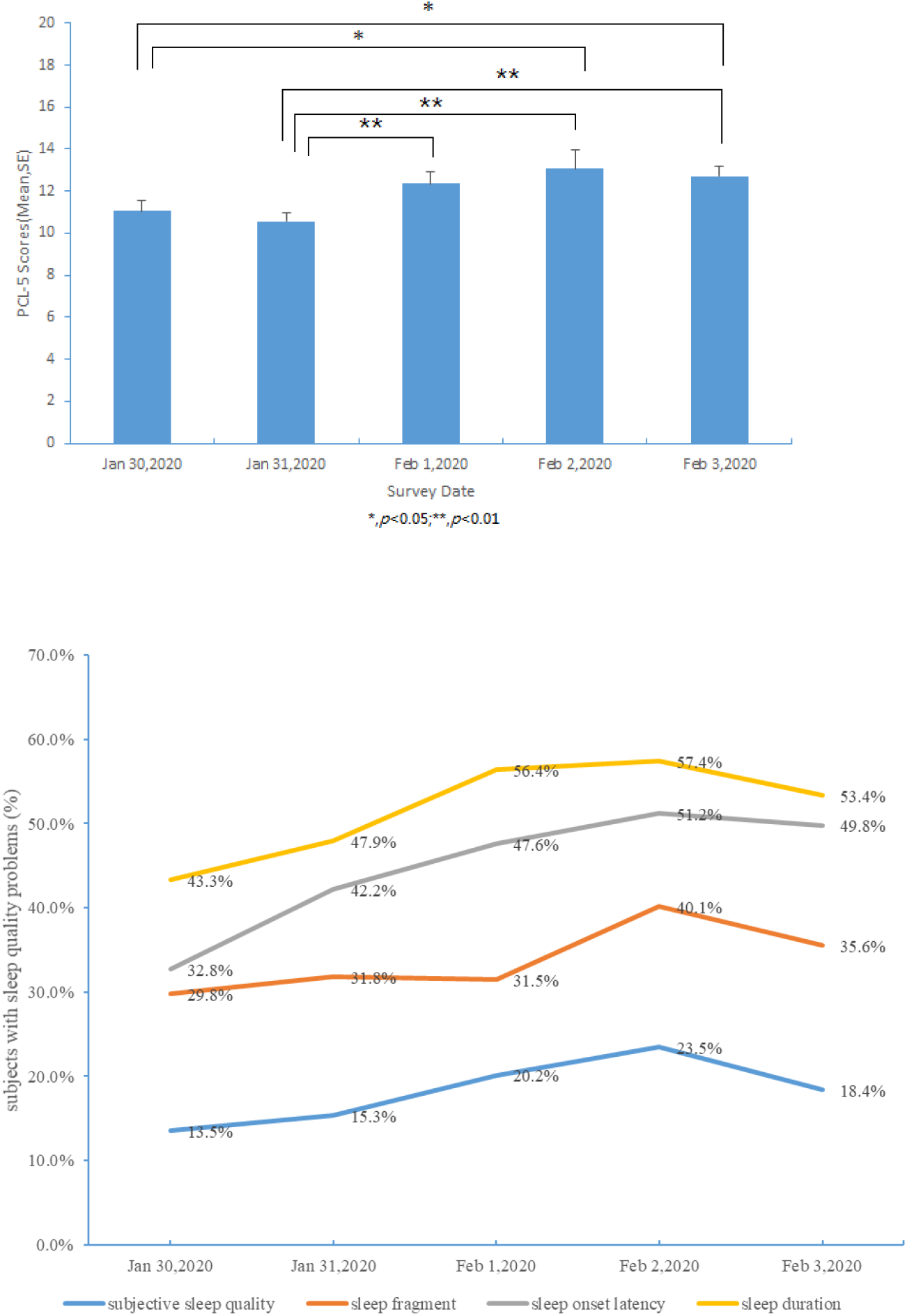
Changes of PCL-5 scores and participants with sleep quality problems in the 5-day survey.

Chi-square test showed significant differences of sleep quality in the 5-days period, χ^*2*^=12.33, *p*<0.05 for subjective sleep quality, χ^*2*^ =34.32, *p*<0.05 for sleep fragment and χ^*2*^=20.03, *p*<0.05 for sleep duration. However, latency onset of sleep did not illustrate significant change during the period.

### Influence of Wuhan exposure history on sleep quality and PTSS

Results demonstrated that participants with Wuhan exposure history generally showed more latency onset of sleep (χ_*2*_=9.77, *p*<0.05) than those not. Wuhan exposure history also significantly influence PCL-5 scores (*t*=-2.93, *p*<0.05). However, there no significant differences of other sleep problems between participants with and without Wuhan-exposure history. See table 2 for detailed information.

**Table 2.** Sleep quality and PCL −5 score by Wuhan exposure history.

| | | Exposure history of Wuhan | | | | $\chi^2$ | $p$-value |
| --- | --- | --- | --- | --- | --- | --- | --- |
|  |  | No(n=1997) |  | Yes(n=230) |  |  |  |
|  |  | N | % | N | % |  |  |
| **Subjective sleep quality** |  |  |  |  |  |  |  |
|  | very good | 724 | 40.3% | 92 | 40.0% | 6.643 | 0.084 |
|  | good | 768 | 42.7% | 91 | 39.6% |  |  |
|  | poor | 281 | 15.6% | 39 | 17.0% |  |  |
|  | very poor | 24 | 1.3% | 8 | 3.5% |  |  |
| **Latency onset of sleep** |  |  |  |  |  |  |  |
|  | No | 1212 | 67.4% | 146 | 63.5% | 9.768 | 0.021 |
|  | <Once a week | 282 | 15.7% | 30 | 13.0% |  |  |
|  | once or twice a week | 218 | 12.1% | 33 | 14.3% |  |  |
|  | >=Three times a week | 85 | 4.7% | 21 | 9.1% |  |  |
| **Sleep fragment** |  |  |  |  |  |  |  |
|  | No | 1001 | 55.7% | 132 | 57.4% | 3.996 | 0.262 |
|  | <Once a week | 286 | 15.9% | 26 | 11.3% |  |  |
|  | once or twice a week | 325 | 18.1% | 43 | 18.7% |  |  |
|  | >=Three times a week | 185 | 10.3% | 29 | 12.6% |  |  |
| **Sleep duration** |  |  |  |  |  |  |  |
|  | >7h | 872 | 48.5% | 124 | 53.9% | 3.861 | 0.277 |
|  | 6-7h | 584 | 32.5% | 61 | 26.5% |  |  |
|  | 5-6h | 276 | 15.4% | 38 | 16.5% |  |  |
|  | <5h | 65 | 3.6% | 7 | 3.0% |  |  |
|  |  | **Mean** | **SD** | **Mean** | **SD** | **$t$** | **$p$-value** |
| **PCL-5 Score** |  | 11.49 | 9.981 | 14.00 | 12.546 | -2.925 | 0.004 |

Note. PCL-5=PTSD Checklist-Civilian Version.

### Interaction of exposure history and sleep quality on PTSS

Table 3 showed results of ANOVAs. To test the exposure history and sleep quality on PTSS, the mean values of PCL-5 entered into 2 × 2 ANOVAs with exposure history and sleep quality as independent variables. Results showed a significant interaction effect of Wuhan exposure history× subjective sleep quality on PCL-5 (*F*=4.01, *p*<0.01, η_p_^2^=0.006). Simple effect tests indicated that there was a significant main effect of Wuhan exposure history on PCL-5 in subjects with relatively poorer subject sleep quality (scored 2 and 3 points in subject sleep quality), but not in subjects with better subject sleep quality. Results also demonstrated interaction of Wuhan exposure history× latency onset of sleep (*F*=3.67, *p*<0.05, η_p_^2^=0.006), but Wuhan-expose only significantly affected PTSS in those showed severe problems falling asleep (score 3 points in latency onset of sleep). Interactions of Wuhan exposure history× sleep fragment (*F*=3.14, *p*<0.05, η_p_^2^=0.006) and Wuhan exposure history× sleep duration (*F*=3.65, *p*<0.05, η_p_^2^=005) were significant. Results of simple effect tests were similar: there were only a main effect of Wuhan exposure history in subjects who were easier to wake up and had shorter sleep duration (scored 3 and 4 points in easily waking during the night or too early in the morning, and in sleep duration). These results indicated that sleep quality regulated the relationship between Wuhan exposure history with sleep and PTSS.

**Table 3.** Interaction of Wuhan exposure history and sleep quality on PCL-5.

| Source | Type III Sum of Squares | df | Mean Square | *F* | *p*-value |
| --- | --- | --- | --- | --- | --- |
| Exposure in Wuhan | 1193.48 | 1 | 1193.48 | 14.321 | <0.001 |
| Subjective sleep quality | 29974.03 | 3 | 9991.34 | 119.891 | <0.001 |
| Exposure in Wuhan × Subjective sleep quality | 1003.67 | 3 | 334.56 | 4.014 | 0.007 |
| Error | 168257.11 | 2019 | 83.34 |  |  |
| Total | 497244.00 | 2027 |  |  |  |
| Exposure in Wuhan | 1007.27 | 1 | 1007.27 | 11.762 | 0.001 |
| Unable to fall asleep within 30 minutes | 26266.76 | 3 | 8755.59 | 102.239 | <0.001 |
| Exposure in Wuhan × Latency onset of sleep | 942.36 | 3 | 314.12 | 3.668 | 0.012 |
| Error | 172903.73 | 2019 | 85.64 |  |  |
| Total | 497244.00 | 2027 |  |  |  |
| Exposure in Wuhan | 1072.77 | 1 | 1072.77 | 12.035 | 0.001 |
| Easily waking during the night or too early in the morning | 19371.99 | 3 | 6457.33 | 72.441 | <0.001 |
| Exposure in Wuhan × Easily waking during the night or too early in the morning | 840.49 | 3 | 280.16 | 3.143 | 0.024 |
| Error | 179972.74 | 2019 | 89.14 |  |  |
| Total | 497244.00 | 2027 |  |  |  |
| Exposure in Wuhan | 1547.88 | 1 | 1547.88 | 15.855 | <0.001 |
| Sleep duration | 10966.04 | 3 | 3655.35 | 37.443 | <0.001 |
| Exposure in Wuhan × Sleep duration | 1070.30 | 3 | 356.77 | 3.654 | 0.012 |
| Error | 197106.05 | 2019 | 97.63 |  |  |
| Total | 497244.00 | 2027 |  |  |  |

## Discussion

According to National Health Commission of China (NHC), until Feb 4, 2020, prevalence rate and fatality rate of COVID-19 in Wuhan were much high above the national level (NHC, 2020), making exposure to Wuhan a risk factor of infection and a cause of mental health problem. The current study aimed to investigate the influence of potential danger of infection on sleep and PTSS. We also explored regulating effects of sleep quality on the relationship between Wuhan exposure history during outbreak of COVID-19 and PTSS. To this end, we conducted ANOVAs and Chi-square tests with the above variables.

Results indicated that data of PTSS growth rapidly in the first 4 days in our study, and decreased in the last day. Interestingly, scores of sleep quality showed the same tendency. This might indicate the close relationship between sleep and PTSD. Moreover, up to the end of our survey, the firmed cases and susceptive cases were still frantically increased, showing no sign of decline (WHO, 2020e). We assumed that results of the present study suggested that emergence of psychological “inflection point” which meant a turning from deterioration of mental health to growth of resilience showed on Feb 3, 2020. It was a date before the number of confirmed cases of COVID-19 showed any sign of downward trends in China. We supposed that the change in psychological response might be a result of Chinese government’s efforts in disease management, psychoeducation and prevention education.

Results illustrated that Wuhan exposure history affected PTSS. For those having Wuhan-expose experiences, PTSS were more obvious. This result was in line with previous findings that subjective perception of dangerousness during SARS related with severity of PTSD symptoms (Mak et al., 2010). Results also highlighted the influence of trauma expose on latency onset of sleep, which was different in subjects with and without Wuhan exposure history. Difficulty in falling asleep might be a result of increased presleep cognitions. According to Fast (2003), traumatic experiences caused reactions of fear, increased arousal and re-experience, and these reactions affected presleep cognitions and leaded to chances of sleep onset latency. Increased sleep onset latency then influenced mood, cognition and physiological responses.

We confirmed that subjective sleep quality, delayed sleep onset, sleep fragment and sleep duration all regulated the effect of Wuhan exposure history on PTSS. Wuhan-expose history influenced only those with relatively poor sleep quality, but if individuals showed few or none sleep problems, Wuhan-expose was not a risk factor of PTSS. These results indicated that traumatic events provide bases for PTSD but one’s ability to keep sleep quality was a protective factor of onset and development of PTSD. This was line with Germain et al.’s (2016) model of sleep and PTSD which indicated that sleep related more closely with PTSS than trauma events. These results were also consist with previous studies which showed alters of REM sleep after uncontrolled stress accounted for the long term physiological and behavioral impairments in an animal model (Vanderheyden et al., 2015). The mechanisms regulating the interactions of traumatic stress and sleep were poorly understood, and we extended previous findings by showing regulation effect of sleep quality on the relationship between trauma expose and PTSD in human samples. To prevent mental illness and protect mental health in the peak rush of COVID-19 and other infectious fatal diseases, health authorities should be mindful of the influence of infectious risks on psychological reactions, propagate the importance of keeping good sleep quality like enough sleep time, keeping early hours etc.

Limitations of the current study included the use of a health example and the fact that we used online survey in data collection. Only a few subjects were confirmed or suspected COVID-19 patients in our study. And the mean levels of sleep quality and PTSS were in the moderate range. For those being infected with COVID-19, sleep and metal health problems might be far more severe. Considering the risk of infection, we used online survey to investigate relationship among Wuhan exposure history, sleep and PTSS. However, it was hard to guarantee authenticity with this method and the validity was limited. We suggested that future studies could exam PTSD and related factors among COVID-19 survivors with face-to-face investigation or interview.

## Conclusion

The present study suggested that sleep quality level decreased and posttraumatic stress symptoms increased with time and change of this tendency was shown on 3^rd^ Feb, 2020. Individuals with and without Wuhan-expose history showed different levels of PTSS and sleep onset latency under the influence of COVID-19. Wuhan-expose history influenced PTSS severity in individuals with poor sleep quality, but not those with high-quality sleep.

## Data Availability

The data used in this study are available from the corresponding author on reasonable request.

